# Effect of image registration on the estimation of pharmacokinetic parameters from DCE-MRI of patients with esophageal cancer

**DOI:** 10.1101/2022.12.17.22283621

**Authors:** Joonsang Lee, Jingfei Ma, Brett Carter, Laurence E. Court, Steven H. Lin

## Abstract

We investigated the effectiveness of the most commonly used registration methods (deformable and rigid-body registrations) with different reference images on pharmacokinetic parameters estimated from dynamic contrast-enhanced magnetic resonance imaging (DCE-MRI) of esophageal cancer patients. We obtained DCE-MRI images from 10 patients with esophageal cancer. Both rigid-body and deformable registrations of the images were performed on DCE-MRI images at different time points as reference images before the pharmacokinetic parameters were estimated. The deformable registration used non-rigid B-spline transforms in a multi-resolution scheme, and Euler transform were used for the rigid body registration. A nonparametric statistical test and the intra-class correlation coefficient assessed the consistency and reproducibility of the pharmacokinetic parameters estimated with both registration methods and using images acquired at different time points. Kruskal-Wallis testing demonstrated significant differences (p < 0.05) in all the estimated parameters for deformable registration but no significant differences (p > 0.78) for rigid-body registration. The intra-class correlation coefficient for rigid-body registration was higher than that for deformable registration for each pharmacokinetic parameter, indicating that, for rigid-body registration, the parameter values from different reference images of one patient tended to be similar to each other. In contrast, the values for deformable registration were more variable. In conclusion, the choice of the reference image of deformable registration significantly affected the estimates of pharmacokinetic parameters, and rigid-body registration showed small variations in pharmacokinetic parameters over the choice of the reference images for small motion artifacts of small distal esophageal cancer on DCE-MRI.

## Introduction

Esophageal cancer is a serious malignant tumor with a high prevalence worldwide ^1^. It is standardly managed with chemoradiation followed by esophagectomy for those patients who can tolerate surgery. Evaluation of the stage and of the response to chemoradiation is very important for the treatment, and several imaging techniques have been used to evaluate esophageal cancer. Nowadays, magnetic resonance imaging (MRI) as a noninvasive and non-radiation technique has been increasingly used in staging and monitoring responses to therapy for esophageal cancer ^2-5^.

MRI plays an important role in cancer diagnosis and treatment planning, providing high-contrast images of soft tissues as well as other useful information based on its applications. One such application is the dynamic contrast-enhanced (DCE)-MRI, which provides information on the microvascular status of a tumor based on the exchange of a contrast agent between the vascular and extravascular/extracellular spaces. DCE-MRI requires repeated acquisition of T1-weighted images of a region of interest (ROI) before, during, and after intravenous administration of a bolus of a paramagnetic contrast agent and provides physiological parameters, such as the volume transfer constant (K^trans^), flux rate constant (kep), extravascular extracellular volume fraction (ve) reflecting vascular permeability ^6-8^, and initial area under the gadolinium curve (IAUGC) ^9^. Applications of DCE-MRI include, among other uses, segmentation of breast carcinomas ^10^, automated deformable image registration ^11^, detection of rheumatoid arthritis ^12^, noninvasive assessment of tumor microenvironments ^13^, and assessment of predictors of clinical outcomes, including treatment response to chemotherapy ^14^.

Because DCE-MRI acquires images of the same region repeatedly over a period of time, respiratory and cardiac motion can result in substantial spatial misregistration between the images from different time points. If uncorrected, such misregistration can affect the required pixel-by-pixel based analysis and accuracy in estimating pharmacokinetic parameters. Although various registration methods are available, rapid change in signal intensity over time makes registration of DCE-MR images challenging, thus limiting the effectiveness of an intensity-based registration algorithm. These rapid intensity changes are not only due to patient motion but also due to the perfusion of the contrast agent in an ROI.

Several registration methods are available for registration of MR images, such as non-rigid-body registration for the kidneys ^15^, deformable and rigid-body registration for prostate cancer ^16^, rigid-body registration for gliomas ^17^, and fixed-frame template-based squared-difference registration for abdominal DCE-MRI data on the liver ^18^. The two most commonly used registration methods with medical images are deformable registration and rigid-body registration. In DCE-MRI, however, there is no standard image registration framework due to relative changes in contrast between two images. In the current study, we sought to determine the effectiveness of both deformable and rigid-body co-registration by applying them to DCE-MRI of patients with esophageal cancer and compared pharmacokinetic parameters estimated using the registered images.

## Materials and Methods

DCE-MRI images of 10 patients with esophageal cancer were examined. This study was approved by the Institutional Review Boards of the University of Texas MD Anderson Cancer Center. All scans were performed using a 3.0T whole-body MRI scanner with an eight-channel torso phased-array coil (GE Healthcare, Waukesha, WI). The DCE-MRI protocol included a standard 3D T1-weighted spoiled gradient echo sequence. The slice thickness was 6 mm, the echo time was 0.80 ms, the repetition time was 2.60 ms, and the number of excitations was one with a 20° flip angle. In total, 120 temporal serial images for each of 24 axial slices were scanned in 13 min.

DCE-MRI images of patients with esophageal cancer were used to compare the pharmacokinetic parameters that were calculated after the DCE-MRI images of the different time points were spatially aligned with rigid-body and deformable registrations. We used the publicly available software program Elastix for the registrations. Elastix uses non-rigid B-spline transforms in a multi-resolution scheme for deformable co-registration and Euler transforms for rigid-body registration. A gradient descent optimizer and mutual information were used as similarity measures for the quality of alignment. In both cases the entire images were used for registration. Table 1 shows the detail parameters in Elastix for deformable registration.

**Table 1.** The detail parameters in Elastix for deformable registration.

| Description | Parameter |
| --- | --- |
| Fixed internal image pixel type | Short |
| Moving internal image pixel type | Short |
| Fixed image dimension | 3 |
| Moving image dimension | 3 |
| Interpolator | B-spline interpolator |
| Resample interpolator | Final B-spline interpolator |
| Fixed image pyramid | Fixed recursive image pyramid |
| Moving image pyramid | Moving recursive image pyramid |
| Optimizer | Adaptive stochastic gradient descent |
| Transform | B-spline transform |
| Final grid spacing in physical units | 40 |
| Number of histogram bins | 32 |
| Number of resolutions | 3 |
| Maximum number of iterations | 500 |
| Number of spatial samples | 5000 |
| B-spline interpolation order | 1 |
| Final B-spline interpolation order | 1 |

In the current study, two images were selected for registration. One was the reference (fixed) image, and the other was the moving image, to be aligned with the reference image. For our testing, three different reference images were selected: a pre-contrast image, an image with maximum uptake of the contrast agent, and a washout image in the tumor. All time serial images used as the moving images were deformed to the reference images, and pharmacokinetic parameters were then calculated for comparison.

The pharmacokinetic analysis of the DCE-MRI data set was performed using the GenIQ software program (GE Healthcare) to estimate pharmacokinetic parameters such as K^trans^, ve, kep, and IAUGC. The slice with the maximum tumor area in each T1-weighted image was selected for this study, and an ROI inside the tumor area was assigned. The number of voxels in the ROIs varied from 81 to 253 (average voxel size in ROIs was 174 ± 81) depending on the tumor sizes on T1-weighted images, and the DCE-MRI data were analyzed on a voxel-by-voxel basis. Fig. 1 shows the signal intensity time curve during the uptake of the contrast agent in an ROI. The reference images of each patient used for co-registration were selected at image frames 10, 20, and 40, which represented the pre-contrast image, image with maximum uptake of the contrast agent, and washout image, respectively. Also, Fig. 2 shows reference MR images of the same patient as in Fig. 1 at these three image frames, as well as a corresponding positron emission tomography image for identifying a tumor location.

**Fig. 1.**
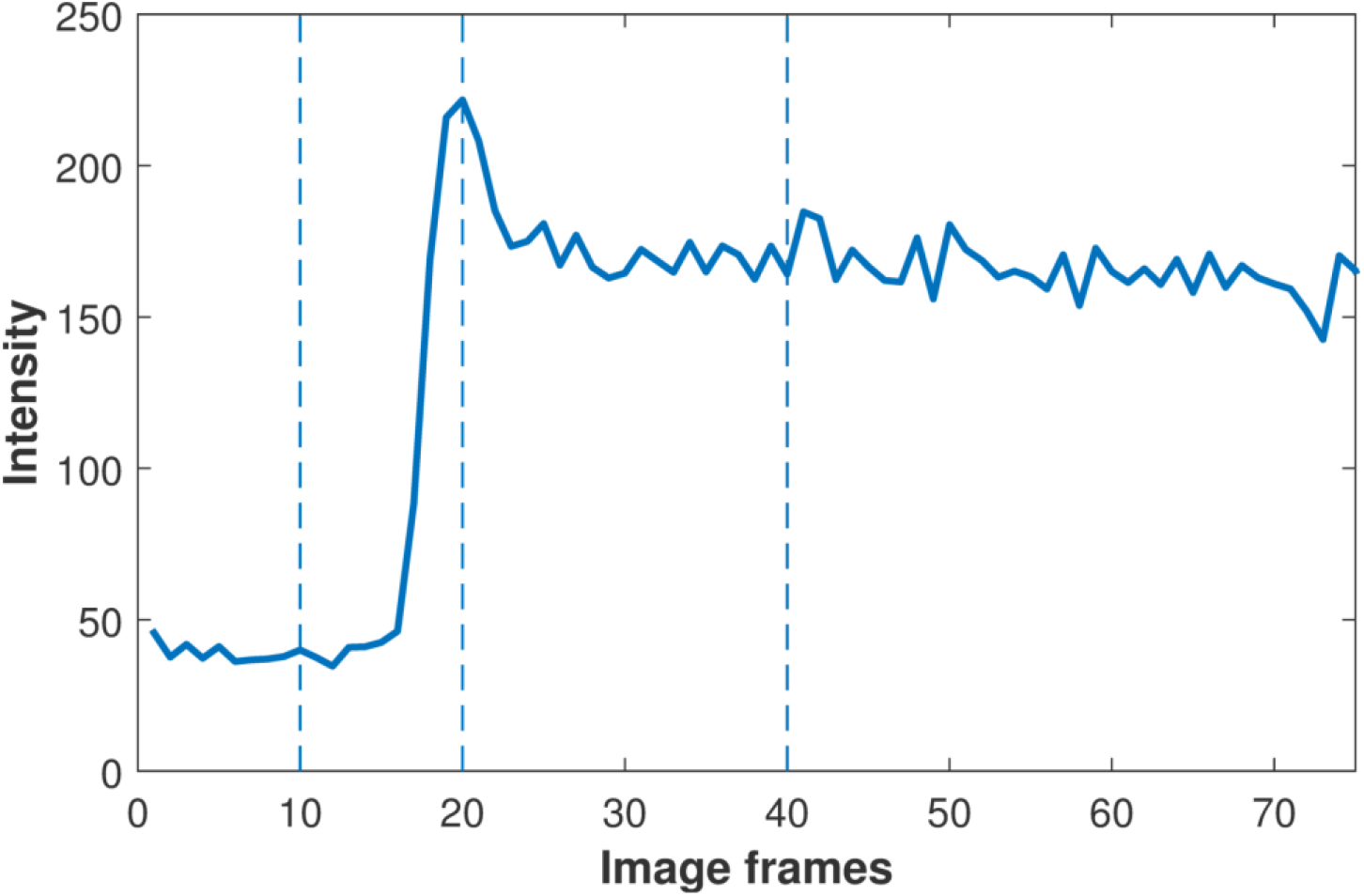
A signal intensity time curve for an ROI in DCE-MRI of an esophageal cancer patient. The images at image frames 10, 20, and 40 were selected as reference images for co-registration.

**Fig. 2.**
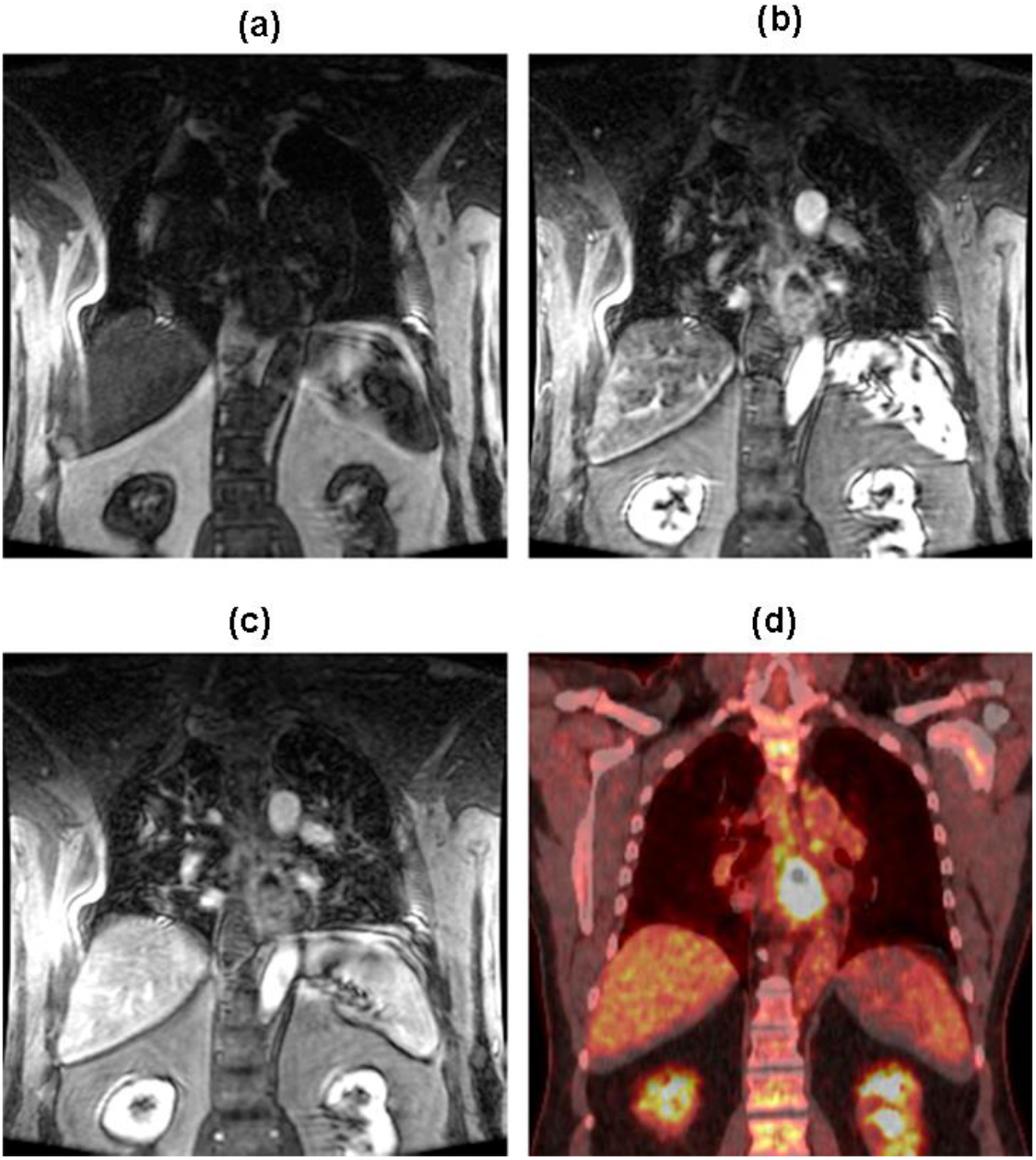
Reference images of each patient for co-registration at image frames 10, 20, and 40 and a corresponding positron emission tomography (PET) image.

### Statistical analysis

The pharmacokinetic parameters K^trans^, ve, kep, and IAUGC were estimated when using each reference image and for both registration methods. For comparison, pharmacokinetic parameters were also estimated using the original MR images without registration. In total, three sets of pharmacokinetic parameters for the two registration methods were obtained for each patient and compared.

The Kruskal-Wallis test, a nonparametric one-way analysis of variance test of the equality of population medians among groups, was used to determine whether the pharmacokinetic parameter values differed significantly between different registration methods. The Kruskal-Wallis test was computed with the following equation

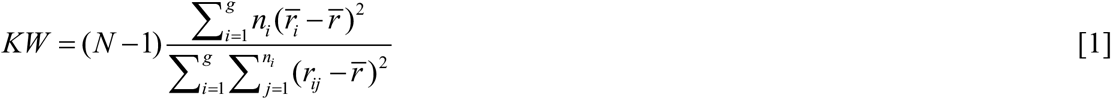

where *n*_*i*_ is the number of measurements in patient *i, r*_*ij*_ is the rank of measurement *j* from patient *i, N* is the total number of measurements across all patients. 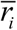 is the average rank of all measurements in patient *i*, and 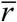 is the average of all the *r*_*ij*_.

Intra-class correlation coefficient (ICC), a measure of the reliability of measurements that can demonstrate how strongly multiple measurements in the same group (patient) resemble each other, was used to assess the consistency and reproducibility of the pharmacokinetic parameters estimated from the different reference images with the two registration methods. Statistical significance was defined as a p value less than 0.05. ICC was computed using the following equation ^19^

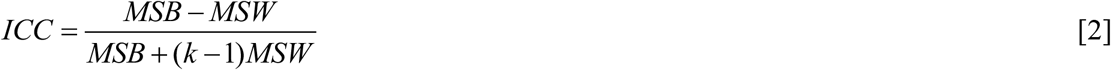

where *MSB* and *MSW* are mean-square-between and mean-square-within, respectively, and *k* is the number of measurements (from three reference images)

## Results

The mean values and standard deviations for K^trans^, v_e_, k_ep_, and IAUGC for the images with deformable registration, rigid registration, or no registration (original images) for all patients are listed in Table 2. In addition, Fig. 3 shows the mean pharmacokinetic parameter values for both registration methods with three different reference images, indicating that the mean pharmacokinetic parameter values for rigid-body registration have less variance than do the values for deformable registration. The blue line in the figure represents a mean pharmacokinetic parameter value for no registration as a reference. Fig. 4 shows the visualization results of both deformable and rigid-body registration. Panels in the top row in the figure 4 (a, b, and c) show the reference images used, and panels d, e, and f show the rigid-body registration.

**Table 2.** Pharmacokinetic parameters estimated after different image co-registration was applied.

| Registration method | Reference image frame | $K^{\text{trans}}$ ($\text{min}^{-1}$) | $k_{\text{ep}}$ ($\text{min}^{-1}$) | $v_e$ | IAUGC ($\text{mmol}\cdot\text{s}^{-1}$) |
| --- | --- | --- | --- | --- | --- |
| None | | $81.72 \pm 25.52$ | $12.91 \pm 2.13$ | $670.36 \pm 133.82$ | $584.36 \pm 136.29$ |
| Deformable | 10 | $86.09 \pm 25.95$ | $18.34 \pm 7.32$ | $519.05 \pm 99.69$ | $524.17 \pm 75.25$ |
| | 20 | $77.34 \pm 23.51$ | $13.74 \pm 2.28$ | $596.99 \pm 131.72$ | $542.66 \pm 137.29$ |
| | 40 | $68.76 \pm 49.72$ | $13.52 \pm 3.53$ | $515.83 \pm 132.43$ | $473.91 \pm 134.19$ |
| Rigid-body | 10 | $82.03 \pm 25.29$ | $12.85 \pm 2.02$ | $675.21 \pm 135.82$ | $587.93 \pm 137.26$ |
| | 20 | $81.72 \pm 25.98$ | $12.76 \pm 1.99$ | $678.09 \pm 136.57$ | $588.28 \pm 142.77$ |
| | 40 | $83.13 \pm 26.35$ | $12.90 \pm 2.08$ | $680.82 \pm 133.63$ | $593.40 \pm 140.17$ |

**Fig. 3.**
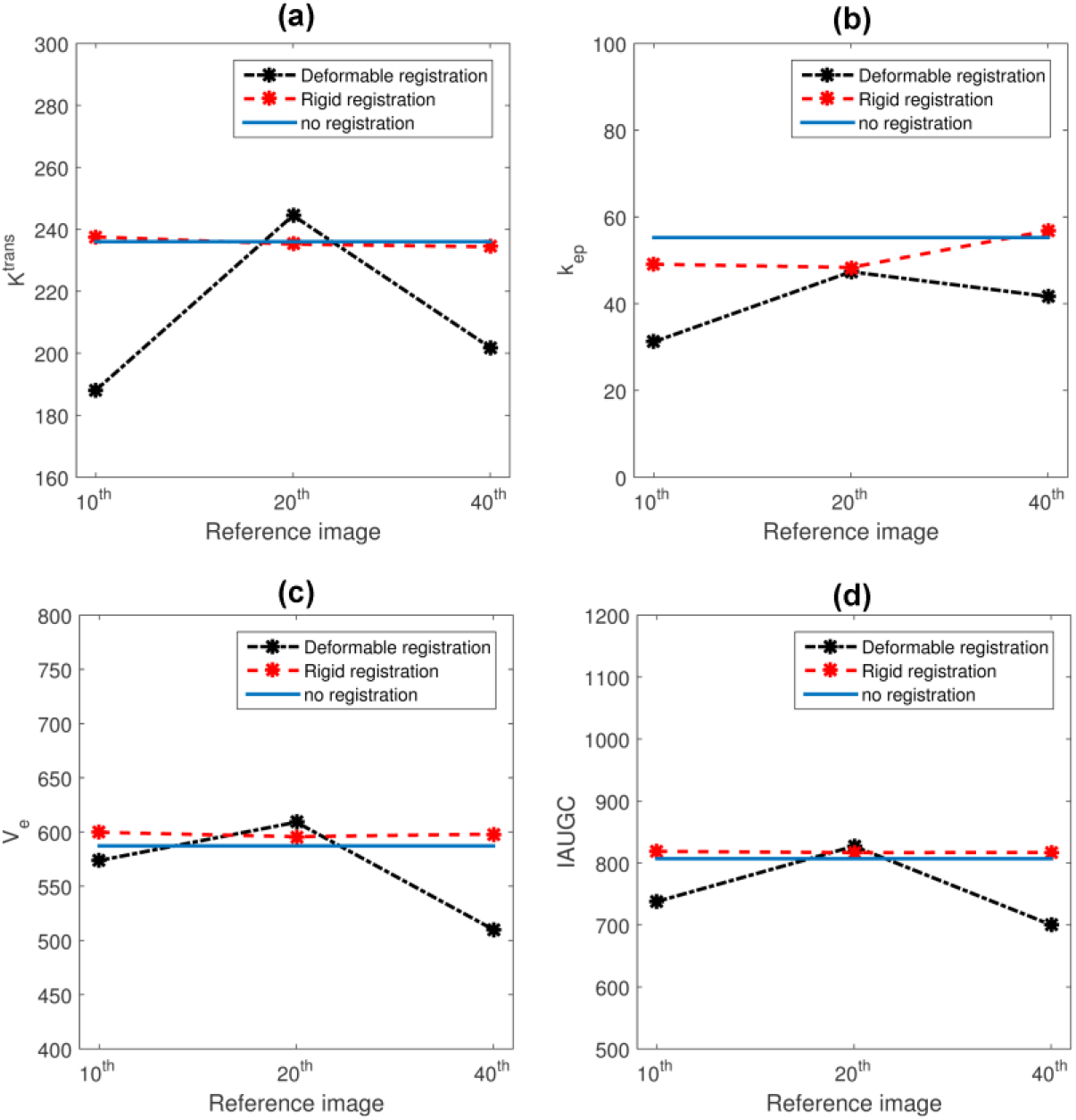
Mean pharmacokinetic parameter values for rigid-body, deformable, and no registration

**Fig. 4.**
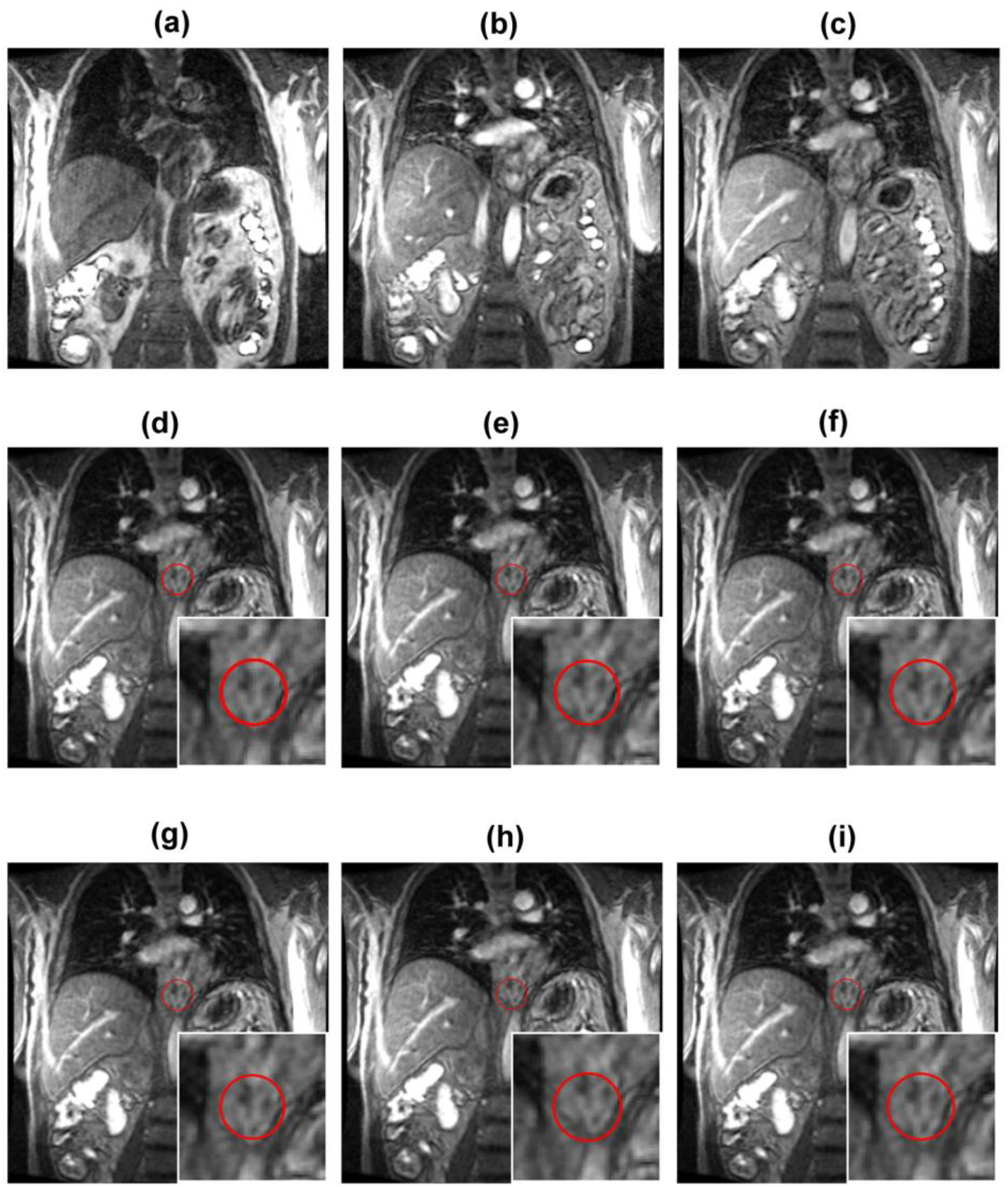
Deformable and rigid-body registration results for a patient. Reference images (a) 10, (b) 20, and (c) 40 were used for both registration. Figure (d) ∼ (f) are the results of the rigid-body registration results and (g) ∼ (i) are the results of the deformable registration.

Kruskal-Wallis testing demonstrated significant differences (p < 0.05) in the pharmacokinetic parameters for three different reference DCE-MR images for deformable registration but no significant differences (p > 0.78) among the groups for rigid registration. The p values for multiple comparisons of the mean ranks for three different reference images with deformable and rigid-body co-registration are listed in Table 3. We used the ICC to assess the consistency and reproducibility of the pharmacokinetic parameters estimated from different reference DCE-MR images with both registration methods. Table 4 shows the ICC for both registration methods, demonstrating how strongly the pharmacokinetic parameters for each reference image in the same patient resembled each other.

**Table 3.** p values (Kruskal-Wallis test) for three different reference DCE-MR images with deformable and rigid-body registration.

| Registration method | $K^{trans}$ | $k_{ep}$ | $v_e$ | IAUGC |
| --- | --- | --- | --- | --- |
| Deformable | $2.98 \times 10^{-12}$ | $1.88 \times 10^{-11}$ | $2.46 \times 10^{-8}$ | $2.31 \times 10^{-7}$ |
| Rigid-body | 0.8828 | 0.7862 | 0.9448 | 0.9302 |

**Table 4.** The ICC for DCE-MRI with deformable and rigid-body registration.

| Pharmacokinetic parameters | ICC |  |
| --- | --- | --- |
|  | Deformable | Rigid-body |
| $K^{trans}$ | 0.8905 | 0.9955 |
| $k_{ep}$ | 0.7306 | 0.9853 |
| $v_e$ | 0.5899 | 0.9994 |
| IAUGC | 0.7829 | 0.9984 |

Fig. 5 shows dot plots of the ICC for both registration methods. We found that the ICC for rigid-body registration was higher (0.9853-0.9994) than that for deformable registration (0.5899-0.8905) for each pharmacokinetic parameter, indicating that rigid-body registration has a relatively lower tendency for values from the same group to be similar than does deformable registration.

**Fig. 5.**
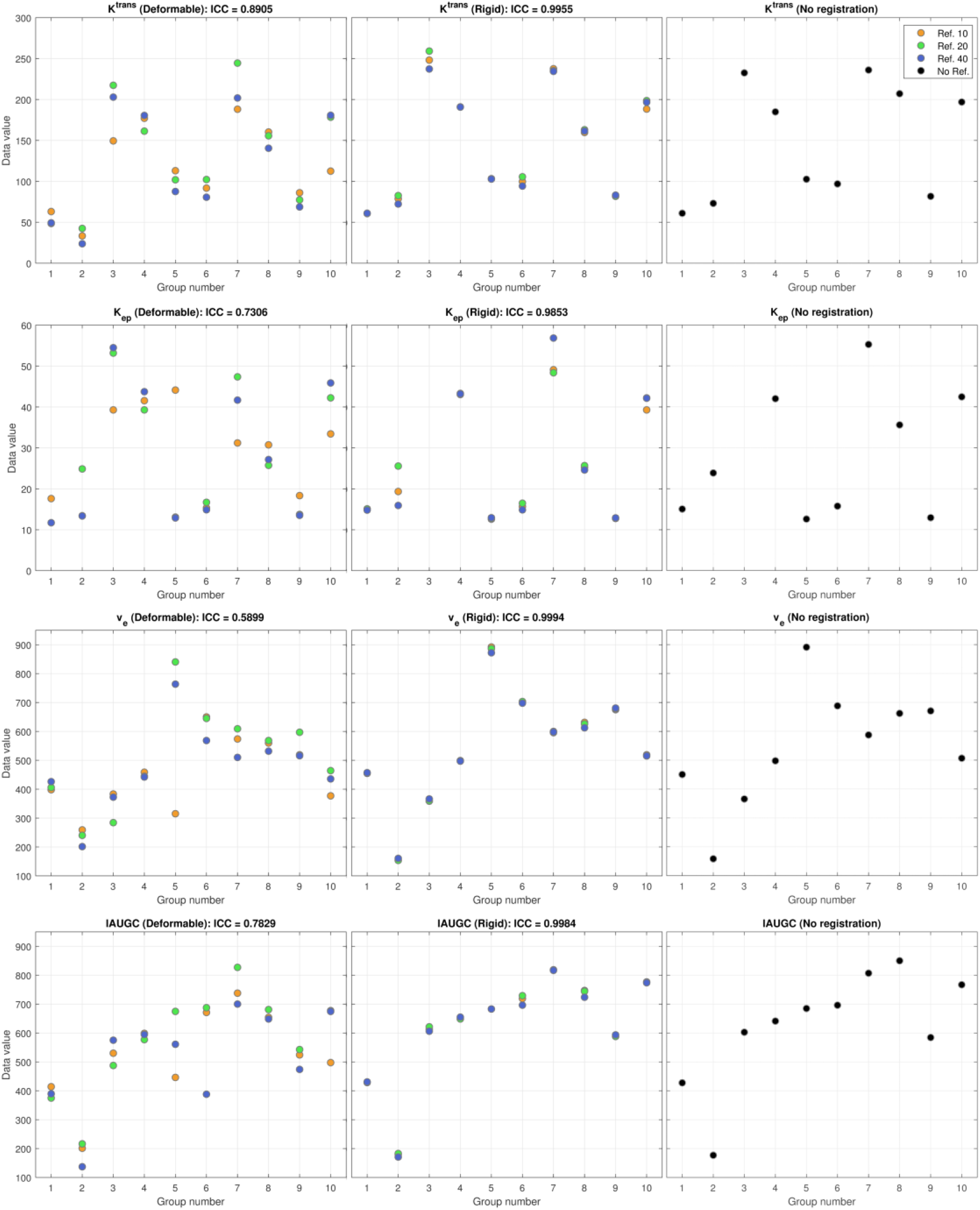
Dot plots of the pharmacokinetic parameter values for the ICCs for DCE-MRI with deformable, rigid-body, and no registration (single pharmacokinetic parameter values for original MR images). In each plot, the x-axis represents patients, and the y-axis represents pharmacokinetic parameter values. In each patient (group), there are three measurements from different reference images for both deformable and rigid-body registration.

## Discussion

We compared the pharmacokinetic parameters estimated from DCE-MRI images of esophageal cancer patients preprocessed with the most commonly used image registration methods: deformable registration with a non-rigid B-spline transform algorithm and rigid-body registration with Euler transform algorithm. Esophageal cancer is usually managed with chemoradiation followed by esophagectomy. Although some esophageal cancer patients (25-30%) do very well after having pathologic complete responses to chemoradiation, most patients have poor responses to treatment and unfavorable prognoses. For those with locally persistent disease after chemoradiation, surgical resection is necessary as salvage therapy. Currently, there are no tools to help clinicians accurately distinguish responders from non-responders among patients with this cancer. In recent studies, researchers investigated multiparametric MRI of esophageal cancer patients that could be helpful for distinguishing chemoradiation responders and non-responders ^4,20-22^.

DCE-MRI is increasingly used in cancer research and has the potential for noninvasively characterizing abnormal microvasculature within a tumor. Specifically, DCE-MRI provides information about blood volume and microvascular permeability with intensity time curves of contrast agent concentrations in dynamic sequential MR images. However, because a DCE-MRI scan takes a long time, motion artifacts such as those caused by patient movement, respiratory motion, and pulsation of the heart cannot be avoided, negatively affecting the accuracy for the estimation of pharmacokinetic parameters.

In DCE-MRI, registration of images can be challenging because the signal intensity in an ROI changes over time. These intensity changes over time are not only due to patient motion or respiratory cycle motion but also due to the perfusion of the contrast agent in a tumor. Intensity enhancement is caused by changes in concentration of the contrast agent, typically a low-molecular-weight gadolinium compound in T1-weighted images, and depends on several tissue physiological factors, such as blood flow, capillary permeability, and interstitial pressure ^23^.

Although many researchers have investigated registration methods with DCE-MRI, a standard image registration framework has yet to be established and the current standard image registration algorithm cannot distinguish intensity changes resulting from patient motion and those caused by perfusion of the contrast agent.

In the current study, we compared the estimated pharmacokinetic parameters after applying the image registration and without the registration. Because we do not have the true pharmacokinetic parameter values for the ROIs as the gold standard, we assumed that we can disregard the motion artifacts in the ROI in the original image for two reasons: the ROI is located close to the center of the field of view and the respiratory cycle motion artifacts in the ROIs were minimal; the average displacements of esophageal tumor in the lateral (LR) and superoinferior (SI) were 0.82 ± 0.55 mm and 2.07 ± 1.42 mm, respectively. This distal esophageal cancer motion due to heart and diaphragm motion agreed with findings of a previous study ^24^. Although the motion artifact is small, this will have an effect on the estimation of the pharmacokinetic parameters on DCE-MRI. Also, we used a mean pharmacokinetic parameter value for no registration as a reference value. This value is not the true value but we can compare the variance of the pharmacokinetic parameter values from multiple measurements between the two registration methods (Fig. 3).

Our results showed that the pharmacokinetic parameter values estimated from the MR images with deformable registration were significantly different (p < 0.05) from those estimated from the MR images with rigid-body registration and those without registration. Furthermore, the pharmacokinetic parameters with the deformable registration using the different reference images differed significantly. In our study, the ICC for each pharmacokinetic parameter was higher with rigid-body registration than with deformable registration. For rigid-body registration, the pharmacokinetic parameter values using the different reference images of a patient were similar to each other, whereas for deformable registration, the parameter values were more varied.

Several researchers have investigated techniques that eliminate the effect of contrast agent for DCE-MRI registration, such as progressive principal component registration (PPCR)^25,26^ and robust principal component analysis (PCA) ^27,28^. However, this is still challenging and has limitations. In our study, we compared the deformable and rigid-body registration methods for small motion artifacts of small distal esophageal cancer on DCE-MRI where the deformation of the ROI due to patient movement artifact is minimized. The rigid-body registration method may help for alignment without affecting intensity changes due to contrast agent in an ROI, especially for small motion artifacts of distal esophageal cancer.

## Conclusions

We investigated the effectiveness and impact of deformable and rigid-body methods of registration of different reference images on the estimated pharmacokinetic parameters of DCE-MRI of patients with distal esophageal cancer. Our results show that the choice of the reference image used for deformable co-registration markedly affected the estimates of the pharmacokinetic parameters.

## Data Availability

All data produced in the present study are available upon reasonable request to the authors.

## Acknowledgements

The authors would like to thank Michael Worley, Donald Norwood, and Dr. Alma Faust for scientific editing. This work was supported by generous support from the Scurlock Foundation to the Center for Radiation Oncology Research at The University of Texas MD Anderson Cancer Center.

## Data Availability

The datasets generated during and/or analyzed during the current study are available from the corresponding author on reasonable request.

## Author Contributions

Project conception and design were by J.L., L.C., J.M., S.L. The data collection and preprocessing were performed by J.L., J.M., B.C. and S.L. The software programming, statistical analysis, and interpretation were performed by J.L. The manuscript was written by J.L. and all authors reviewed the manuscript.

